# SARS-CoV-2 Infection is Associated with an Increase in New Diagnoses of Schizophrenia Spectrum and Psychotic Disorder: A Study Using the US National COVID Cohort Collaborative (N3C)

**DOI:** 10.1101/2023.12.05.23299473

**Authors:** Asif Rahman, Michael Russell, Wanhong Zheng, Daniel Eckrich, Imtiaz Ahmed, the N3C Consortium

## Abstract

Amid the ongoing global repercussions of SARS-CoV-2, it’s crucial to comprehend its potential long-term psychiatric effects. Several recent studies have suggested a link between COVID-19 and subsequent mental health disorders. Our investigation joins this exploration, concentrating on Schizophrenia Spectrum and Psychotic Disorders (SSPD). Different from other studies, we took acute respiratory distress syndrome (ARDS) and COVID-19 lab negative cohorts as control groups to accurately gauge the impact of COVID-19 on SSPD. Data from 19,344,698 patients, sourced from the N3C Data Enclave platform, were methodically filtered to create propensity matched cohorts: ARDS (n = 222,337), COVID-positive (n = 219,264), and COVID-negative (n = 213,183). We systematically analyzed the hazard rate of new-onset SSPD across three distinct time intervals: 0-21 days, 22-90 days, and beyond 90 days post-infection. COVID-19 positive patients consistently exhibited a heightened hazard ratio (HR) across all intervals [0-21 days (HR: 4.6; CI: 3.7-5.7), 22-90 days (HR: 2.9; CI: 2.3 -3.8), beyond 90 days (HR: 1.7; CI: 1.5-1.)]. These are notably higher than both ARDS and COVID-19 lab-negative patients. Validations using various tests, including the Cochran Mantel Haenszel Test, Wald Test, and Log-rank Test confirmed these associations. Intriguingly, our data indicated that younger individuals face a heightened risk of SSPD after contracting COVID-19, a trend not observed in the ARDS and COVID-negative groups. These results, aligned with the known neurotropism of SARS-CoV-2 and earlier studies, accentuate the need for vigilant psychiatric assessment and support in the era of Long-COVID, especially among younger populations.

## Introduction

It has been over three years since the initial identification of SARS-CoV-2 infection (hereafter referred to as COVID-19)in the USA. Despite the development of a vaccine and efforts to combat the pandemic, there are still many unanswered questions. Particularly, the long-term effects of COVID on mental health are yet to be fully unwrapped and associated with the disease. Several preliminary studies [1–3] have suggested an increased risk of mental illness following a COVID-19 diagnosis, including but not limited to anxiety, depression, mood disorder, post-traumatic stress disorder (PTSD), insomnia, dementia, delirium, encephalitis, psychosis, and nerve disorder. It is important to note that viral infections resulting from recent outbreaks of severe acute respiratory syndrome (SARS) in 2002 and Middle East respiratory syndrome (MERS) in 2012, both caused by coronavirus closely related to SARS-CoV-2, were also associated with neurological manifestations in some cases [3].

COVID-19 has a multi-organ pathology that includes the human brain and the central nervous system [4]. It has been detected both in the brain and cerebrospinal fluids of the diagnosed patients. The COVID-19 patients show a greater cognitive decline compared to the non-COVID patients. It has also been associated with brain structural change [5]. Recent studies have suggested that more than one-third of the infected individuals develop neurological symptoms in the acute phase of the disease, and around 34% of them show brain abnormalities [6, 7]. COVID-19 has been linked to excessive and dysregulated immune responses that can lead to systemic inflammation. Patients with severe COVID-19 have been found to have elevated levels of various inflammatory markers in their blood, such as C-reactive protein (CRP), ferritin, interleukin-6 (IL-6), and others. Among them, higher levels of CRP and IL-6 are linked to a higher risk of different neurological conditions such as major depressive disorders.

It has been well established that a bidirectional interaction exists between the central nervous system, specifically the brain, and systemic inflammation [9]. In the brain, microglia are the principal cells involved in modulating the effects of remote inflammatory stressors resulting in neuro-inflammatory manifestations of systemic processes arising from multiple causes (trauma, infection, auto-immune processes, etc.) [10]. Microglial functional and structural alterations have been found in multiple major psychiatric disorders [10–12] although presence, degree and configuration of such alterations vary between diagnoses. Therefor the etiologic and therapeutic significance of these observations remains unclear [13].

A growing body of literature has implicated systemic inflammation associated with critical illness in the development of delirium [17]. In turn, the occurrence of delirium during critical illness is associated with persistent deficits in neurocognitive function following survival [18]. The presence of pre-exposure decline in cognitive function is associated with an increased risk of post-critical illness persistent neurocognitive disability [19, 20]. Difference in epigenetic DNA methylation patterns in critically ill patients with and without delirium have recently been reported [21]. It is now well established that systemic inflammation affects brain function in critical illness and that these effects are persistent beyond the intensive care episode [22, 23]. While the exact mechanisms are still incompletely characterized, epigenetic modification of DNA directed protein transcription may play a potential role.

Major psychiatric illness is known to have a strong genetic component. However, variable penetrance suggests that environmental factors are also important in the development of clinical disease [24]. Emerging data suggests a significant association between neuroinflammatory changes and major psychiatric illness. The strongest associations to date involve the development of schizophrenia, bipolar disorder and major depression [10, 11, 25].

Given the increasing evidence demonstrating a link between systemic inflammation and neurocognitive function, along with the role of neuroinflammation in manifesting major psychiatric disorders and the systemic inflammatory effects of COVID-19, the authors sought to establish if COVID-19 could lead to a rise in the onset of significant psychiatric conditions. We decide to concentrate on the schizophrenia spectrum and psychotic disorder (SSPD), as the data linking inflammatory conditions with the emergence or progression of the disease is most compelling in this context.

We are aware that several studies [2, 3, 8] tried to establish an association between COVID-19 and psychiatric manifestations. However, a significant portion of these studies lacked an appropriate comparison group, leading to an incomplete understanding of the incidence and prevalence of neuro-psychiatric disorders in COVID-19 patients. To address this limitation, our study incorporated a control group comprising individuals with Acute Respiratory Distress Syndrome (ARDS) and those who tested negative for COVID-19. This approach ensured a more precise correlation between COVID-19 and SSPD. Furthermore, we leveraged the N3C platform, utilizing its vast, robust, and long-term data set to effectively discern and quantify the impact of COVID-19 on SSPD.

## Materials and Methods

This study was a retrospective cohort study. All data was collected from the National COVID-19 Cohort Collaborative (N3C) Data Enclave platform. The dataset was retrieved on May 31, 2023, and we limited our analysis to include only records up to that date. Throughout the process of data collection and subsequent analysis, the authors did not have access to any information that could be used to identify individual participants.

### Study Design and Data Collection

To achieve our objectives, we initiated a systematic filtering process as depicted in Figure 1. Out of an initial dataset of 19 million patients, we categorized them into three primary groups: Acute Respiratory Distress Syndrome (ARDS), COVID-positive, and COVID-negative. We applied specific criteria to refine these cohorts. Firstly, we considered only those with a minimum of three visits. Secondly, we excluded patients with any pre-existing mental health conditions and further narrowed down our scope to individuals aged between 17 and 70 years. Notably, within the COVID-positive group, we focused on patients characterized by moderate, severe, or terminal outcomes due to the virus. After implementing these filters, our COVID-positive cohort size was finalized at 244,226. To ensure that our data from these groups could be directly compared, we also implemented a propensity score matching technique.

**Fig 1.**
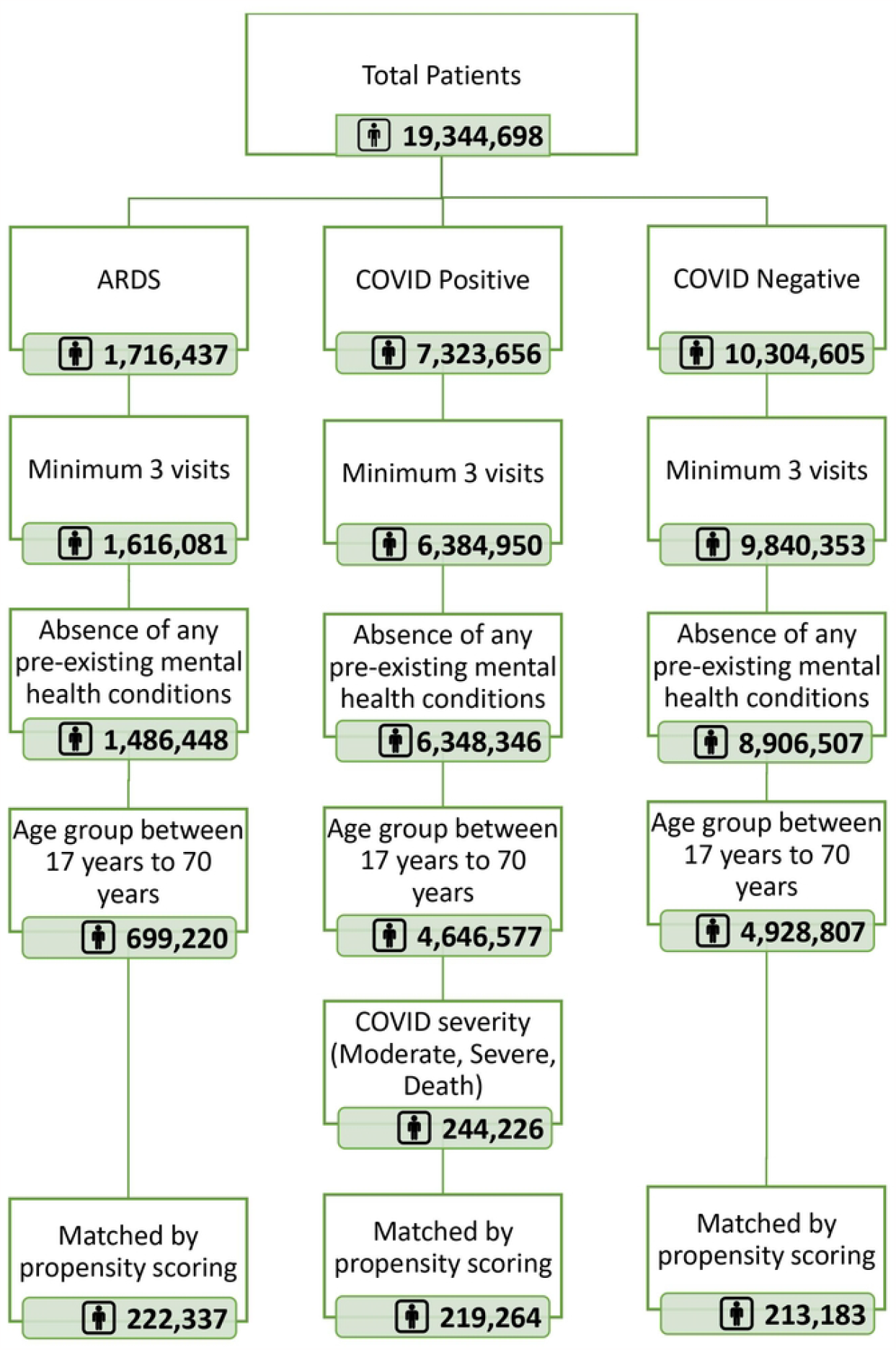
Final cohort selection by applying exclusion criteria

### Cohorts

Three distinct cohorts were constructed for this study: one case and two controls. The case cohort was made up of patients diagnosed with COVID-19, based on the N3C defined computable phenotype version 4.0. [50]. In order to derive meaningful insights, we limited our COVID-19 positive patients to those with moderate to severe manifestations. This categorization was determined by several factors, including the duration of inpatient hospital stays, usage of invasive ventilation, application of extracorporeal membrane oxygenation (ECMO), and even unfortunate fatalities.

For our controls, we selected patients diagnosed with ARDS post-January 1st, 2020 but without any record of a COVID-19 diagnosis during the pandemic. The third group consisted of those who tested negative for COVID-19 and had no prior history of either COVID-19 or ARDS. The starting population for our study needed to have a history of at least three medical visits spanning 365 days or more. To standardize the timeline across cohorts, index dates were determined based on the earliest date of relevant diagnosis or lab test results.

We excluded individuals below 17 years or over 70 years at the time of the index date. Furthermore, patients with the following mental health disorders prior to the index date were also removed from consideration:

- Schizophrenia Spectrum Disorders
- Bipolar Disorders
- Major Depression
- Personality Disorders
- Trauma

For homogeneity, all cohorts underwent 1:1 nearest neighbor (NN) matching on propensity scores using the R “MatchIt” package. This matching was based on the following attributes:

- Gender
- Race
- Ethnicity
- Age groups
- Prior psychiatric drug prescription or administration
- Prior Hypothyroidism diagnosis
- Prior Anxiety diagnosis
- Prior Substance Abuse diagnosis
- Prior Insomnia diagnosis

Please refer to the appendix for specific Observational Medical Outcomes Partnership (OMOP) codesets [51] used for identifying these attributes. Post-matching, the cohort sizes were 219,264 for COVID-19 positive patients, 213,183 for the lab negatives, and 222,337 for ARDS patients.

### Outcome

Once the cohorts were built, we looked at the first incident post-index date of SSPD. The code sets were developed using the OMOP concept and concept ancestor tables [51] and reviewed by subject matter experts. While psychiatry continued to debate over the relationship between psychotic symptoms and mood symptoms [52, 53], in this study, to focus on the hallmark thought symptoms characteristic of schizophrenia (such as delusions, hallucinations, or disorganized speech), we separated SSPD from bipolar and depressive disorders. Therefore, while acute psychotic disorder, schizophreniform, schizophrenia, schizoaffective disorders, and related ICD-10 diagnoses were included in the SSPD category, mood disorders with psychotic features were counted in the latter two categories and thus excluded from the analysis.

The data was subsequently structured to include three pivotal columns: a boolean flag denoting whether a patient was diagnosed with SSPD after the index date, the exact date of the patient’s initial SSPD diagnosis post the index date, and the duration in days between the index date and this diagnosis. This data arrangement culminated in a matrix with a single record for each patient, encompassing cohort identifiers, demographic details, other relevant covariates, and the critical outcome variable.

### Statistical Analysis

To examine the association between COVID-19 and SSPD, we performed a comparative analysis among the matched cohorts using the N3C platform in R (version 4.0). Our primary predictor variable is the disease type, categorized into three groups: COVID-positive, COVID-negative, and ARDS. The outcome variable is binary, indicating either SSPD or non-SSPD.

We utilized the Cox Proportional Hazard Model [43] to derive the hazard ratios (HR) of COVID-positive patients relative to the COVID-negative and ARDS patients. The time-to-event for patients diagnosed with SSPD was measured from their SSPD diagnosis date up to the COVID and ARDS reference dates, which include the dates of their positive COVID test, negative COVID lab test, and ARDS diagnosis. For patients without an SSPD diagnosis, this duration was taken from their most recent recorded visit to the reference date of either their COVID or ARDS diagnosis.

Before deploying the Cox model, it was imperative to test the proportional hazard assumption. In doing so, the Schoenfeld residual analysis, a conventional diagnostic tool for this purpose [44], yielded a significant p-value. This necessitated rejecting the null hypothesis of a uniform proportional hazard over the comprehensive time frame of 180 days. Consequently, we segmented the cohort into three distinct time intervals: 0-21 days, 22-90 days, and beyond 90 days. These intervals were subsequently validated for the proportional hazard assumptions, and the Cox model was then applied to each to ascertain the hazard ratio (HR).

In tandem with the Cox model, we also conducted the Cochran Mantel Haenszel Test [45], the Likelihood Ratio Test [46], the Wald Test [45], and the Log-rank Test [47] across the three time intervals. A p-value threshold of 0.05 served as the determinant for statistical significance in all these tests.

## Results

The results shed light on the potential long-term psychiatric implications of SARS-CoV-2 infection. The study found robust evidence linking SARS-CoV-2 infection to an augmented risk of Schizophrenia Spectrum and Psychotic Disorders (SSPD). Specifically, COVID-positive patients displayed almost double the incident rate (0.56%) compared to COVID-negative (0.33%) and ARDS (0.29%) patients, as showcased in Table 1.

**Table 1.** Characteristics of Cohorts.

| Characteristics | ARDS |  | COVID Positive |  | COVID Negative |  |
| --- | --- | --- | --- | --- | --- | --- |
|  | N = 222337 |  | N = 219264 |  | N = 213183 |  |
|  | Count | Percentage | Count | Percentage | Count | Percentage |
| **Gender:** |  |  |  |  |  |  |
| Female | 105175 | 47.30% | 103528 | 47.20% | 105959 | 49.70% |
| Male | 117162 | 52.70% | 115736 | 52.80% | 107196 | 50.30% |
| Other or Unknown | 0 | 0.00% | 0 | 0.00% | 28 | 0.01% |
| **Age Groups:** |  |  |  |  |  |  |
| 21 and Younger | 7896 | 3.55% | 7449 | 3.40% | 7840 | 3.68% |
| 22 to 29 | 19885 | 8.94% | 19160 | 8.74% | 19644 | 9.21% |
| 30 to 39 | 31070 | 14.00% | 30298 | 13.80% | 29400 | 13.80% |
| 40 to 49 | 35871 | 16.10% | 34407 | 15.70% | 31594 | 14.80% |
| 50 to 59 | 52529 | 23.60% | 53059 | 24.20% | 51115 | 24.00% |
| 60 and Older | 75086 | 33.80% | 74891 | 34.20% | 73590 | 34.50% |
| **Race:** |  |  |  |  |  |  |
| Asian | 5519 | 2.48% | 5580 | 2.54% | 5568 | 2.61% |
| Black/African American | 48251 | 21.70% | 47047 | 21.50% | 45269 | 21.20% |
| White/Caucasian | 132914 | 59.80% | 126061 | 57.50% | 127741 | 59.90% |
| Unknown | 29357 | 13.20% | 31810 | 14.50% | 27014 | 12.70% |
| Nothing mentioned | 1746 | 0.79% | 2701 | 1.23% | 2758 | 1.29% |
| Other | 4550 | 2.05% | 6065 | 2.77% | 4833 | 2.27% |
| **Ethnicity:** |  |  |  |  |  |  |
| Hispanic or Latino | 34320 | 15.40% | 38196 | 17.40% | 31002 | 14.50% |
| Not Hispanic or Latino | 171609 | 77.20% | 165071 | 75.30% | 166149 | 77.90% |
| Missing/Unknown | 16408 | 7.38% | 15997 | 7.30% | 16032 | 7.52% |
| **Any Psychiatric Disease (before index date):** |  |  |  |  |  |  |
| No | 145612 | 65.50% | 138029 | 63.00% | 137490 | 64.50% |
| Yes | 76725 | 34.50% | 81235 | 37.00% | 75693 | 35.50% |
| **Anxiety Disorder:** |  |  |  |  |  |  |
| No | 198285 | 89.20% | 201933 | 92.10% | 192958 | 90.50% |
| Yes | 24052 | 10.80% | 17331 | 7.90% | 20225 | 9.49% |
| **Insomnia Disorder:** |  |  |  |  |  |  |
| No | 211921 | 95.30% | 212449 | 96.90% | 204869 | 96.10% |
| Yes | 10416 | 4.68% | 6815 | 3.11% | 8314 | 3.90% |
| **Hypothyroidism Disorder:** |  |  |  |  |  |  |
| No | 206762 | 93.00% | 204850 | 93.40% | 198391 | 93.10% |
| Yes | 15575 | 7.01% | 14414 | 6.57% | 14792 | 6.94% |
| **Hypothyroidism Disorder (Lab Test):** |  |  |  |  |  |  |
| No | 212801 | 95.70% | 208228 | 95.00% | 201818 | 94.70% |
| Yes | 9536 | 4.29% | 11036 | 5.03% | 11365 | 5.33% |
| **Drug Abuse / Addiction Disorder:** |  |  |  |  |  |  |
| No | 189594 | 85.30% | 190323 | 86.80% | 188360 | 88.40% |
| Yes | 32743 | 14.70% | 28941 | 13.20% | 24823 | 11.60% |
| **Drug Abuse / Addiction Disorder (Lab Test):** |  |  |  |  |  |  |
| No | 218686 | 98.40% | 211424 | 96.40% | 209335 | 98.20% |
| Yes | 3651 | 1.64% | 7840 | 3.58% | 3848 | 1.81% |
| **All Mental Health:** |  |  |  |  |  |  |
| No | 207448 | 93.30% | 203123 | 92.60% | 197204 | 92.50% |
| Yes | 14889 | 6.70% | 16141 | 7.36% | 15979 | 7.50% |
| **SSPD:** |  |  |  |  |  |  |
| No | 221700 | 99.70% | 218035 | 99.40% | 212476 | 99.70% |
| Yes | 637 | 0.29% | 1229 | 0.56% | 707 | 0.33% |
| **Bipolar Disorder:** |  |  |  |  |  |  |
| No | 214506 | 96.50% | 210281 | 95.90% | 205014 | 96.20% |
| Yes | 7831 | 3.52% | 8983 | 4.10% | 8169 | 3.83% |
| **Personality Disorder:** |  |  |  |  |  |  |
| No | 222034 | 99.90% | 218951 | 99.90% | 212889 | 99.90% |
| Yes | 303 | 0.14% | 313 | 0.14% | 294 | 0.14% |
| **Depression:** |  |  |  |  |  |  |
| No | 210088 | 94.50% | 205253 | 93.60% | 199865 | 93.80% |
| Yes | 12249 | 5.51% | 14011 | 6.39% | 13318 | 6.25% |
| **Trauma:** |  |  |  |  |  |  |
| No | 219432 | 98.70% | 217107 | 99.00% | 210308 | 98.70% |
| Yes | 2905 | 1.31% | 2157 | 0.98% | 2875 | 1.35% |

Using the Cox model, a marked difference in the hazard ratio of new-onset psychiatric outcomes became evident between COVID-19 positive patients and the cohorts of ARDS and COVID-19 lab negative patients (Please refer to Table 2). For all time intervals considered, COVID-negative patients were the benchmark for hazard ratio computations. In the immediate 21 days following exposure, the hazard ratio for COVID-positive patients was notably high (HR: 4.6; 95% CI: 3.7 to 5.7) when contrasted with ARDS patients (HR: 0.73 CI: 0.53 to 0.99). This suggests that, during the acute phase, individuals positive for COVID-19 were significantly more likely to be diagnosed with SSPD than both their COVID-negative and ARDS counterparts.

**Table 2.** Hazard Ratios with 95% Confidence Interval (CI) For SSPD In Different Time Frames.

| Time Frame (Days) | COVID Negative |  |  |  | COVID Positive |  |  |  | ARDS |  |  |
| --- | --- | --- | --- | --- | --- | --- | --- | --- | --- | --- | --- |
|  | Hazard Ratio | 95 % CI |  |  | Hazard Ratio | 95 % CI |  |  | Hazard Ratio | 95 % CI |  |
|  |  | Lower | Upper |  |  | Lower | Upper |  |  | Lower | Upper |
| 0 - 21 | * | * | * |  | 4.6 | 3.7 | 5.7 |  | 0.73 | 0.53 | 0.99 |
| 22 - 90 | * | * | * |  | 2.9 | 2.3 | 3.8 |  | 1.1 | 0.79 | 1.43 |
| Beyond 90 | * | * | * |  | 1.7 | 1.5 | 1.9 |  | 0.97 | 0.86 | 1.47 |

\*COVID Negative patient group is considered as references.

In the subsequent interval of 22-90 days, the hazard ratio for COVID-positive patients remained elevated (HR: 2.9; 95% CI: 2.3 to 3.8), and interestingly, ARDS patients exhibited their peak hazard ratio of the study (HR:1.1, CI: 0.79 to 1.43). Beyond 90 days, the hazard ratio for the COVID-positive group experienced a reduction (HR: 1.7; 95% CI: 1.5 to 1.9) yet was consistently higher than that of the ARDS patients (HR:0.97, CI: 0.86 to 1.47). Despite the reduction in hazard ratios as time progressed, it’s salient to note that COVID-19 survivors remain at a heightened risk for SSPD well beyond the immediate aftermath of their infection. The hazard ratios for different time intervals are visually summarized in Figure 2.

**Fig. 2.**
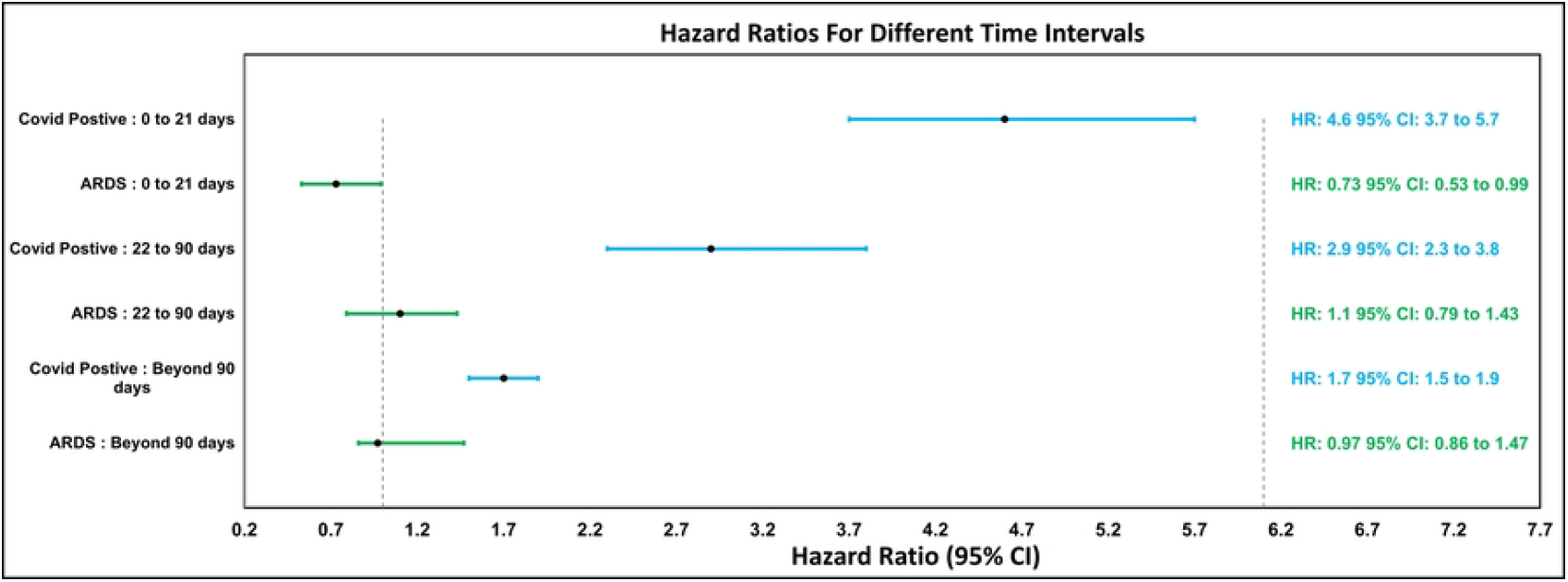
SSPD Hazard Ratio Comparisons.

Following the detailed hazard ratio analysis, several statistical tests (Please refer to Table 3) were conducted to further validate the findings. The Schoenfeld residuals rest returned a p-value exceeding 0.05, suggesting the retention of the null hypothesis that the Hazard Ratio is consistent over time. Additionally, tests such as the Cochran Mantel Haenszel Test, Likelihood Ratio Test, Wald Test, and Log-rank Test consistently showed p-values less than 0.05, reinforcing a significant association between being COVID-positive and receiving an SSPD diagnosis.

**Table 3.** P Value For Different Tests In Different Time Frames.

| P Value | Time Frame (In Days) |  |  |
| --- | --- | --- | --- |
|  | 0 - 21 | 22 - 90 | Beyond 90 |
| Schoenfeld Residuals Test | 0.24 | 0.75 | 0.15 |
| Cochran Mantel Haenszel Test | < 0.05 | < 0.05 | < 0.05 |
| Likelihood Ratio Test | < 0.05 | < 0.05 | < 0.05 |
| Wald Test | < 0.05 | < 0.05 | < 0.05 |
| Logrank Test | < 0.05 | < 0.05 | < 0.05 |

In our efforts to determine potential demographic factors influencing SSPD occurrence among COVID-positive patients, several key demographics emerged as more susceptible to SSPD following a COVID-19 diagnosis. Specifically, males, individuals aged 21 or younger, those of African American descent, and non-Hispanic or Latino individuals showcased a heightened vulnerability (see Table 4 and Table 5 in the appendix for details). Intriguingly, these same demographic trends, with the exception of the age factor, were mirrored in the SSPD occurrence among both the COVID-negative and ARDS groups (refer to Table 6 and Table 7 in the appendix for further insights). While these findings undoubtedly warrant a deeper exploration, we strongly advocate for future research endeavors to prioritize this critical observation regarding the younger people being more susceptible to post-COVID SSPD.

**Table 4.** SSPD Patient Comparison Among ARDS, COVID Positive and COVID Negative Group.

|  | ARDS Patient N = 637 |  | COVID Positive Patient N =1229 |  | COVID Negative Patient = 707 |  | P Value |
| --- | --- | --- | --- | --- | --- | --- | --- |
|  | Count | Percentage | Count | Percentage | Count | Percentage |  |
| **Gender:** |  |  |  |  |  |  |  |
| Female | 222 | 35% | 518 | 42% | 244 | 35% | <2.2e-16 |
| Male | 415 | 65% | 711 | 58% | 463 | 65% | <2.2e-16 |
| **Age Groups:** | 637 | Total | 1229 | Total | 707 | Total |  |
| 21 and Younger | # | # | 62 | 5.04% | 28 | 3.96% | 4.91E-8 |
| 22 to 29 | <65 | <10.20% | 112 | 9.11% | 73 | 10.33% | 0.0001715 |
| 30 to 39 | 94 | 14.76% | 181 | 14.73% | 116 | 16.41% | 1.52E-07 |
| 40 to 49 | 113 | 17.74% | 170 | 13.83% | 112 | 15.84% | 0.0002312 |
| 50 to 59 | 164 | 25.75% | 263 | 21.40% | 156 | 22.07% | 1.15E-08 |
| 60 and Older | 190 | 29.83% | 441 | 35.88% | 222 | 31.40% | <2.2e-16 |
| **Race:** | 637 | Total | 1229 | Total | 707 | Total |  |
| Black/African American | 246 | 38.62% | 361 | 29.37% | 279 | 39.46% | 6.98E-06 |
| Asian and White/Caucasian | 297 | 46.62% | 711 | 57.85% | 319 | 45.12% | <2.2e-16 |
| **Ethnicity:** |  |  |  |  |  |  |  |
| Hispanic or Latino | 73 | 11.46% | 118 | 9.60% | 88 | 12.45% | 0.003535 |
| Not Hispanic or Latino | 507 | 79.59% | 1015 | 82.59% | 571 | 80.76% | <2.2e-16 |

The (#) marked group has a count or proportion that is too small to quantitatively report

**Table 5.**
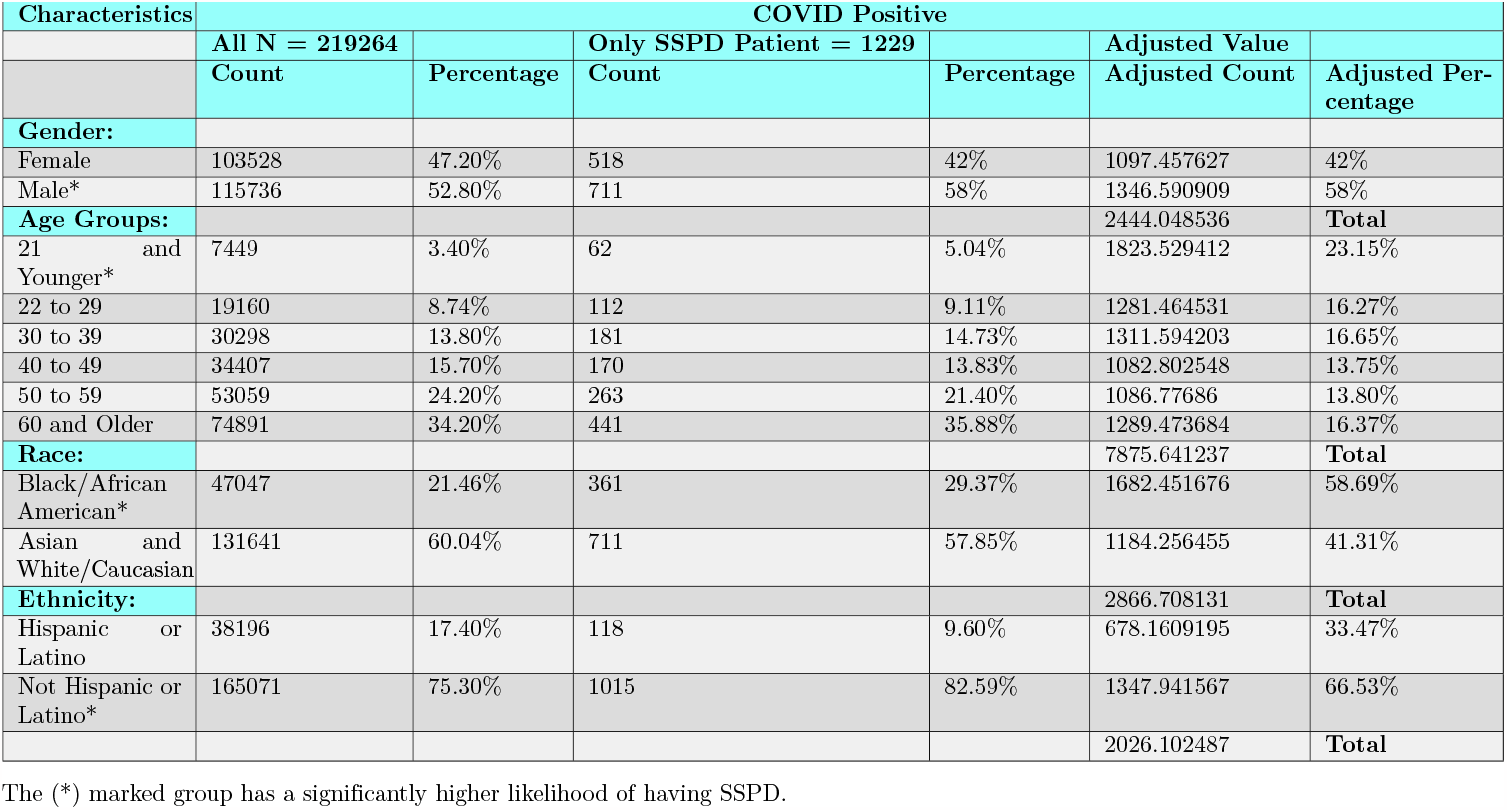
SSPD Patient Comparison Within COVID Positive Group.

**Table 6.**
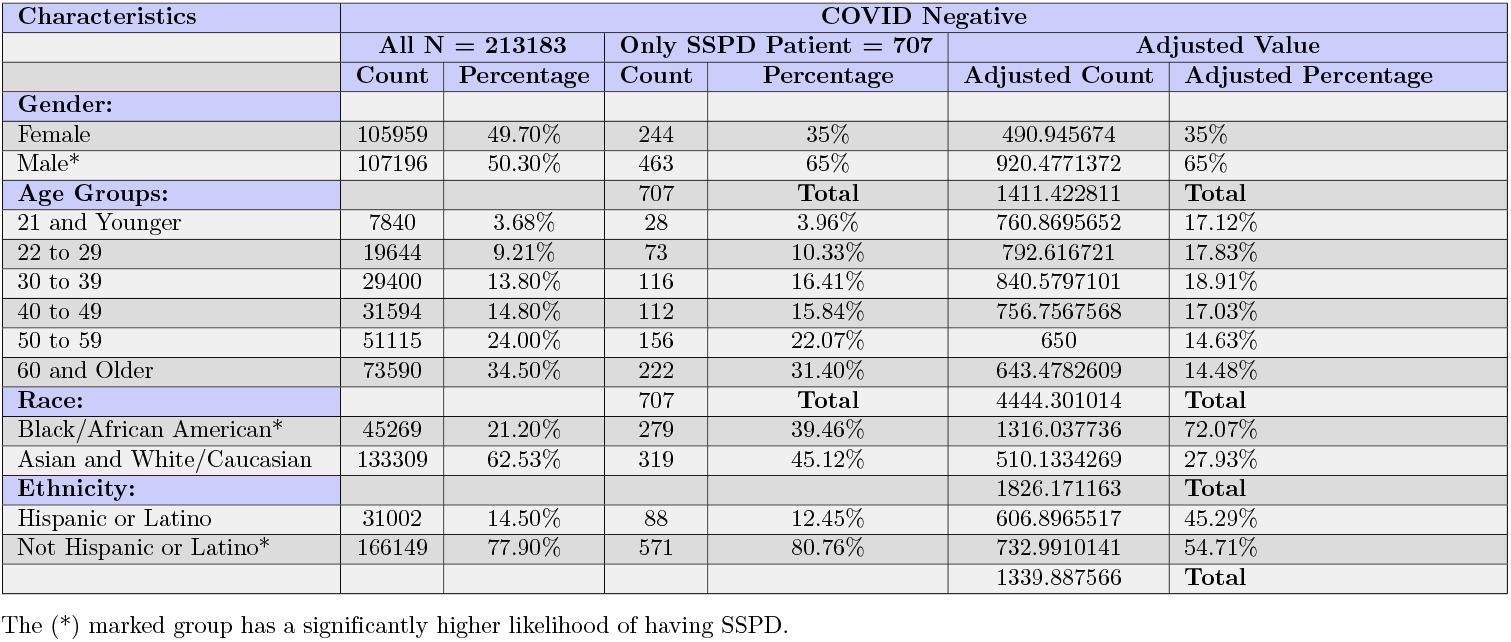
SSPD Patient Comparison Within COVID Negative Group.

**Table 7.**
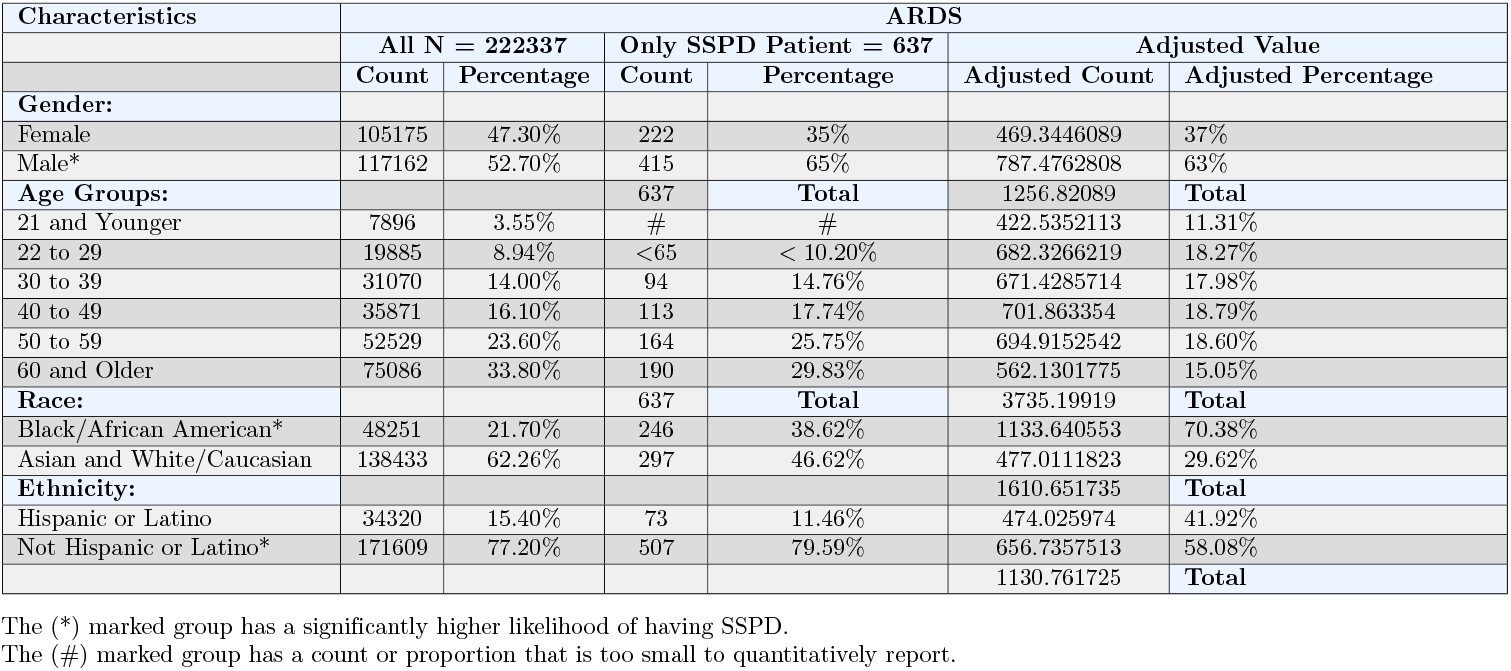
SSPD Patient Comparison Within ARDS Group.

## Discussion

Our study indicates that the likelihood of developing SSPD after a COVID-19 infection is higher than in ARDS and COVID-19-negative patients. The significance of various demographic factors has also emerged from our results. These insights underscore the vital importance of keeping a close watch on the mental well-being of those recovering from COVID-19. Their persistent increased risk points to a wider societal concern, especially regarding severe psychiatric conditions like SSPD.

Extensive literature has accumulated since 2000 that indicates an association between various inflammatory markers, changes in the structure and function of a variety of cellular components of the brain, and the development of major psychiatric illnesses. No longer thought to be “immunologically privileged” by virtue of the blood-brain permeability barrier, it is now well established that the brain is extensively influenced by systemic inflammation and, in turn, can modulate systemic inflammation through descending [26] and biochemical [27] pathways. Inflammatory influences in the brain are structural [28] and functional [9]. Effects of maternal development on in-utero brain development and subsequent offspring behavior have been demonstrated in animal models [29] although the relevance to mental illness in humans remains to be determined. Inflammatory influences on the developed brain have also been demonstrated and tied to clinically relevant behavior [15, 30]. However, studies evaluating the relationship between various inflammatory markers and clinically relevant behavior have yielded inconsistent results [17, 32–34]. At the moment, these inflammatory changes cannot be causally linked directly to specific conditions, but they do provide potential mechanistic insights and may become plausible therapeutic targets in conditions where the response to currently available medications varies widely [17, 28, 30].

With that background, our group sought to establish whether any association existed between SARS-CoV-2 infection and the extensive accompanying inflammatory response and new-onset psychiatric illness. Our results are consistent with the hypothesis that COVID-19 infection and the accompanying inflammatory state (“cytokine storm”) is positively associated with the new onset of SSPD. Further, these results strongly suggest a direct relationship between the development of SSPD and the severity of the disease state and presumably the intensity of the attendant inflammatory response.

The strengths of our study are underscored by the utilization of an expansive and meticulously curated national dataset, enabling an in-depth analysis of a vast number of records. Furthermore, our rigorous approach to defining exclusionary criteria and cohort matching bolster the robustness of our findings. However, the study does possess notable limitations. Foremost among these is, same as any other retrospective studies, the dependence on documented diagnoses (ICD-10 coding) to pinpoint new instances of SSPD. It is conceivable — perhaps even probable — that certain patients might have been inaccurately diagnosed, thereby skewing their categorization as determined by our data extraction methodology. The markedly elevated hazard ratio observed during the acute (0-21 day) phase, for instance, is challenging to rationalize, given that definitive diagnoses for many severe psychiatric conditions, especially SSPD, typically necessitate prolonged periods of behavioral observation. One possible reason might be that many individuals went through an early phase of unnoticed or undeclared symptoms before their clinical visit for COVID. When diagnosing SSPD, doctors likely factored in this extended course of symptoms, likely triggered by the brain inflammation caused by the coronavirus infection. There also exists a possibility that some individuals could have encountered acute and transient psychotic disorders (ATPD), which were erroneously identified as SSPD. Additionally, patients manifesting early signs hinting at SSPD may have exhibited more distinct symptoms post-COVID infection, subsequently leading to accurate diagnostic coding. Absent direct clinical assessments, pinpointing the primary influencing factor remains elusive. Nonetheless, the sustained elevated hazard ratios even beyond the initial 90 days post-infection underscore the continued influence of COVID-19 on the emergence of SSPD.

## Conclusion

In this study, we have found a substantial increase in the likelihood of being diagnosed with a schizophrenia spectrum and psychotic disorder (SSPD) after experiencing moderate to severe illness due to SARS-CoV-2 infection, in comparison to a group of individuals who had non-COVID Acute Respiratory Distress Syndrome (ARDS). Our work is consistent with the known neurotropism of the SARS-CoV-2 virus [39, 40] and other reports of increased risk of major psychiatric disorders following COVID-19 infection [41–43]. Further research is required to identify specific characteristics of populations and individuals who may be at a particularly high risk of developing SSPD and potentially other significant psychiatric conditions following COVID-19 infection. Understanding these psychiatric risks associated with COVID-19 is an essential component of our strategy to address the evolving landscape of Long-COVID.

## Data Availability

The data underlying the results presented in the study are available from https://covid.cd2h.org/ which requires institutional access.

## Acknowledgments

We would like to acknowledge West Virginia University (WVU) and WVU Clinical & Translational Research Institute (CTSI) for their support. The project described was supported by the National Institute Of General Medical Sciences, 5U54GM104942-04. The content is solely the responsibility of the authors and does not necessarily represent the official views of the NIH.

## N3C Attribution

The analyses described in this publication were conducted with data or tools accessed through the NCATS N3C Data Enclave (https://covid.cd2h.org) and N3C Attribution & Publication Policy v 1.2-2020-08-25b supported by NCATS U24 TR002306. This research was possible because of the patients whose information is included within the data and the organizations (https://ncats.nih.gov/n3c/resources/data-contribution/data-transfer-agreement-signatories) and scientists who have contributed to the ongoing development of this community resource [https://doi.org/10.1093/jamia/ocaa196].

## Disclaimer

The N3C Publication Committee confirmed that this manuscript MSID:1645.616 is in accordance with N3C data use and attribution policies; however, this content is solely the responsibility of the authors and does not necessarily represent the official views of the National Institutes of Health or the N3C program.

## IRB

The N3C data transfer to NCATS is performed under a Johns Hopkins University Reliance Protocol # IRB00249128 or individual site agreements with NIH. The N3C Data Enclave is managed under the authority of the NIH; information can be found at https://ncats.nih.gov/n3c/resources.

## Individual Acknowledgements For Core Contributors

We gratefully acknowledge the following core contributors to N3C: Adam B. Wilcox, Adam M. Lee, Alexis Graves, Alfred (Jerrod) Anzalone, Amin Manna, Amit Saha, Amy Olex, Andrea Zhou, Andrew E. Williams, Andrew Southerland, Andrew T. Girvin, Anita Walden, Anjali A. Sharathkumar, Benjamin Amor, Benjamin Bates, Brian Hendricks, Brijesh Patel, Caleb Alexander, Carolyn Bramante, Cavin Ward-Caviness, Charisse Madlock-Brown, Christine Suver, Christopher Chute, Christopher Dillon, Chunlei Wu, Clare Schmitt, Cliff Takemoto, Dan Housman, Davera Gabriel, David A. Eichmann, Diego Mazzotti, Don Brown, Eilis Boudreau, Elaine Hill, Elizabeth Zampino, Emily Carlson Marti, Emily R. Pfaff, Evan French, Farrukh M Koraishy, Federico Mariona, Fred Prior, George Sokos, Greg Martin, Harold Lehmann, Heidi Spratt, Hemalkumar Mehta, Hongfang Liu, Hythem Sidky, J.W. Awori Hayanga, Jami Pincavitch, Jaylyn Clark, Jeremy Richard Harper, Jessica Islam, Jin Ge, Joel Gagnier, Joel H. Saltz, Joel Saltz, Johanna Loomba, John Buse, Jomol Mathew, Joni L. Rutter, Julie A. McMurry, Justin Guinney, Justin Starren, Karen Crowley, Katie Rebecca Bradwell, Kellie M. Walters, Ken Wilkins, Kenneth R. Gersing, Kenrick Dwain Cato, Kimberly Murray, Kristin Kostka, Lavance Northington, Lee Allan Pyles, Leonie Misquitta, Lesley Cottrell, Lili Portilla, Mariam Deacy, Mark M. Bissell, Marshall Clark, Mary Emmett, Mary Morrison Saltz, Matvey B. Palchuk, Melissa A. Haendel, Meredith Adams, Meredith Temple-O’Connor, Michael G. Kurilla, Michele Morris, Nabeel Qureshi, Nasia Safdar, Nicole Garbarini, Noha Sharafeldin, Ofer Sadan, Patricia A. Francis, Penny Wung Burgoon, Peter Robinson, Philip R.O. Payne, Rafael Fuentes, Randeep Jawa, Rebecca Erwin-Cohen, Rena Patel, Richard A. Moffitt, Richard L. Zhu, Rishi Kamaleswaran, Robert Hurley, Robert T. Miller, Saiju Pyarajan, Sam G. Michael, Samuel Bozzette, Sandeep Mallipattu, Satyanarayana Vedula, Scott Chapman, Shawn T. O’Neil, Soko Setoguchi, Stephanie S. Hong, Steve Johnson, Tellen D. Bennett, Tiffany Callahan, Umit Topaloglu, Usman Sheikh, Valery Gordon, Vignesh Subbian, Warren A. Kibbe, Wenndy Hernandez, Will Beasley, Will Cooper, William Hillegass, Xiaohan Tanner Zhang. Details of contributions are available at https://covid.cd2h.org/core-contributors

## Data Partners with Released Data

The following institutions whose data is released or pending: Available: Advocate Health Care Network — UL1TR002389: The Institute for Translational Medicine (ITM) • Aurora Health Care Inc — UL1TR002373: Wisconsin Network For Health Research • Boston University Medical Campus — UL1TR001430: Boston University Clinical and Translational Science Institute • Brown University — U54GM115677: Advance Clinical Translational Research (Advance-CTR) • Carilion Clinic — UL1TR003015: iTHRIV Integrated Translational health Research Institute of Virginia • Case Western Reserve University — UL1TR002548: The Clinical & Translational Science Collaborative of Cleveland (CTSC) • Charleston Area Medical Center — U54GM104942: West Virginia Clinical and Translational Science Institute (WVCTSI) • Children’s Hospital Colorado — UL1TR002535: Colorado Clinical and Translational Sciences Institute • Columbia University Irving Medical Center — UL1TR001873: Irving Institute for Clinical and Translational Research • Dartmouth College — None (Voluntary) Duke University — UL1TR002553: Duke Clinical and Translational Science Institute • George Washington Children’s Research Institute — UL1TR001876: Clinical and Translational Science Institute at Children’s National (CTSA-CN) • George Washington University — UL1TR001876: Clinical and Translational Science Institute at Children’s National (CTSA-CN) • Harvard Medical School — UL1TR002541: Harvard Catalyst • Indiana University School of Medicine — UL1TR002529: Indiana Clinical and Translational Science Institute • Johns Hopkins University — UL1TR003098: Johns Hopkins Institute for Clinical and Translational Research • Louisiana Public Health Institute — None (Voluntary) • Loyola Medicine — Loyola University Medical Center • Loyola University Medical Center — UL1TR002389: The Institute for Translational Medicine (ITM) • Maine Medical Center — U54GM115516: Northern New England Clinical & Translational Research (NNE-CTR) Network • Mary Hitchcock Memorial Hospital & Dartmouth Hitchcock Clinic — None (Voluntary) • Massachusetts General Brigham — UL1TR002541: Harvard Catalyst • Mayo Clinic Rochester — UL1TR002377: Mayo Clinic Center for Clinical and Translational Science (CCaTS) • Medical University of South Carolina — UL1TR001450: South Carolina Clinical & Translational Research Institute (SCTR) • MITRE Corporation — None (Voluntary) • Montefiore Medical Center — UL1TR002556: Institute for Clinical and Translational Research at Einstein and Montefiore • Nemours — U54GM104941: Delaware CTR ACCEL Program • NorthShore University HealthSystem — UL1TR002389: The Institute for Translational Medicine (ITM) • Northwestern University at Chicago — UL1TR001422: Northwestern University Clinical and Translational Science Institute (NUCATS) • OCHIN — INV-018455: Bill and Melinda Gates Foundation grant to Sage Bionetworks • Oregon Health & Science University — UL1TR002369: Oregon Clinical and Translational Research Institute • Penn State Health Milton S. Hershey Medical Center — UL1TR002014: Penn State Clinical and Translational Science Institute • Rush University Medical Center — UL1TR002389: The Institute for Translational Medicine (ITM) • Rutgers, The State University of New Jersey — UL1TR003017: New Jersey Alliance for Clinical and Translational Science • Stony Brook University — U24TR002306 • The Alliance at the University of Puerto Rico, Medical Sciences Campus — U54GM133807: Hispanic Alliance for Clinical and Translational Research (The Alliance) • The Ohio State University — UL1TR002733: Center for Clinical and Translational Science • The State University of New York at Buffalo — UL1TR001412: Clinical and Translational Science Institute • The University of Chicago — UL1TR002389: The Institute for Translational Medicine (ITM) • The University of Iowa — UL1TR002537: Institute for Clinical and Translational Science • The University of Miami Leonard M. Miller School of Medicine — UL1TR002736: University of Miami Clinical and Translational Science Institute • The University of Michigan at Ann Arbor — UL1TR002240: Michigan Institute for Clinical and Health Research • The University of Texas Health Science Center at Houston — UL1TR003167: Center for Clinical and Translational Sciences (CCTS) • The University of Texas Medical Branch at Galveston — UL1TR001439: The Institute for Translational Sciences • The University of Utah — UL1TR002538: Uhealth Center for Clinical and Translational Science • Tufts Medical Center — UL1TR002544: Tufts Clinical and Translational Science Institute • Tulane University — UL1TR003096: Center for Clinical and Translational Science • The Queens Medical Center — None (Voluntary) • University Medical Center New Orleans — U54GM104940: Louisiana Clinical and Translational Science (LA CaTS) Center • University of Alabama at Birmingham — UL1TR003096: Center for Clinical and Translational Science • University of Arkansas for Medical Sciences — UL1TR003107: UAMS Translational Research Institute • University of Cincinnati — UL1TR001425: Center for Clinical and Translational Science and Training • University of Colorado Denver, Anschutz Medical Campus — UL1TR002535: Colorado Clinical and Translational Sciences Institute • University of Illinois at Chicago — UL1TR002003: UIC Center for Clinical and Translational Science • University of Kansas Medical Center — UL1TR002366: Frontiers: University of Kansas Clinical and Translational Science Institute • University of Kentucky — UL1TR001998: UK Center for Clinical and Translational Science • University of Massachusetts Medical School Worcester — UL1TR001453: The UMass Center for Clinical and Translational Science (UMCCTS) • University Medical Center of Southern Nevada — None (voluntary) • University of Minnesota — UL1TR002494: Clinical and Translational Science Institute • University of Mississippi Medical Center — U54GM115428: Mississippi Center for Clinical and Translational Research (CCTR) • University of Nebraska Medical Center — U54GM115458: Great Plains IDeA-Clinical & Translational Research • University of North Carolina at Chapel Hill — UL1TR002489: North Carolina Translational and Clinical Science Institute • University of Oklahoma Health Sciences Center — U54GM104938: Oklahoma Clinical and Translational Science Institute (OCTSI) • University of Pittsburgh — UL1TR001857: The Clinical and Translational Science Institute (CTSI) • University of Pennsylvania — UL1TR001878: Institute for Translational Medicine and Therapeutics • University of Rochester — UL1TR002001: UR Clinical & Translational Science Institute • University of Southern California — UL1TR001855: The Southern California Clinical and Translational Science Institute (SC CTSI) • University of Vermont — U54GM115516: Northern New England Clinical & Translational Research (NNE-CTR) Network • University of Virginia — UL1TR003015: iTHRIV Integrated Translational health Research Institute of Virginia • University of Washington — UL1TR002319: Institute of Translational Health Sciences • University of Wisconsin-Madison — UL1TR002373: UW Institute for Clinical and Translational Research • Vanderbilt University Medical Center — UL1TR002243: Vanderbilt Institute for Clinical and Translational Research • Virginia Commonwealth University — UL1TR002649: C. Kenneth and Dianne Wright Center for Clinical and Translational Research • Wake Forest University Health Sciences — UL1TR001420: Wake Forest Clinical and Translational Science Institute • Washington University in St. Louis — UL1TR002345: Institute of Clinical and Translational Sciences • Weill Medical College of Cornell University — UL1TR002384: Weill Cornell Medicine Clinical and Translational Science Center • West Virginia University — U54GM104942: West Virginia Clinical and Translational Science Institute (WVCTSI)• Submitted: Icahn School of Medicine at Mount Sinai — UL1TR001433: ConduITS Institute for Translational Sciences • The University of Texas Health Science Center at Tyler — UL1TR003167: Center for Clinical and Translational Sciences (CCTS) • University of California, Davis — UL1TR001860: UCDavis Health Clinical and Translational Science Center • University of California, Irvine — UL1TR001414: The UC Irvine Institute for Clinical and Translational Science (ICTS) • University of California, Los Angeles — UL1TR001881: UCLA Clinical Translational Science Institute • University of California, San Diego — UL1TR001442: Altman Clinical and Translational Research Institute • University of California, San Francisco — UL1TR001872: UCSF Clinical and Translational Science Institute • Pending: Arkansas Children’s Hospital — UL1TR003107: UAMS Translational Research Institute • Baylor College of Medicine — None (Voluntary) • Children’s Hospital of Philadelphia — UL1TR001878: Institute for Translational Medicine and Therapeutics • Cincinnati Children’s Hospital Medical Center — UL1TR001425: Center for Clinical and Translational Science and Training • Emory University — UL1TR002378: Georgia Clinical and Translational Science Alliance • HonorHealth — None (Voluntary) • Loyola University Chicago — UL1TR002389: The Institute for Translational Medicine (ITM) • Medical College of Wisconsin — UL1TR001436: Clinical and Translational Science Institute of Southeast Wisconsin • MedStar Health Research Institute — None (Voluntary) • Georgetown University — UL1TR001409: The Georgetown-Howard Universities Center for Clinical and Translational Science (GHUCCTS) • MetroHealth — None (Voluntary) • Montana State University — U54GM115371: American Indian/Alaska Native CTR • NYU Langone Medical Center — UL1TR001445: Langone Health’s Clinical and Translational Science Institute • Ochsner Medical Center — U54GM104940: Louisiana Clinical and Translational Science (LA CaTS) Center • Regenstrief Institute — UL1TR002529: Indiana Clinical and Translational Science Institute • Sanford Research — None (Voluntary) • Stanford University — UL1TR003142: Spectrum: The Stanford Center for Clinical and Translational Research and Education • The Rockefeller University — UL1TR001866: Center for Clinical and Translational Science • The Scripps Research Institute — UL1TR002550: Scripps Research Translational Institute • University of Florida — UL1TR001427: UF Clinical and Translational Science Institute • University of New Mexico Health Sciences Center — UL1TR001449: University of New Mexico Clinical and Translational Science Center • University of Texas Health Science Center at San Antonio — UL1TR002645: Institute for Integration of Medicine and Science • Yale New Haven Hospital — UL1TR001863: Yale Center for Clinical Investigation.

## A Appendix A: Tables in Appendix

## B Appendix B: Supporting Information

**S1 Table** Schizophrenia Spectrum and Psychotic Disorders Code Set: list of diagnosis codes used to evaluate outcomes

**S2 Table** Exclusion Code Set: list of diagnosis codes used to exclude patients if one or more codes were included in the patient record prior to the index date.

**S3 Table** Matching Criteria Code Set: diagnosis and drug exposure codes used in the R MatchIt package to build case and control cohorts.

